# Follow-up of cancer incidence associated with smoke-related PM_2.5_ exposure due to a coal mine fire

**DOI:** 10.1101/2024.11.13.24317068

**Authors:** Sunav N Nayagam, Pei Yu, Caroline X Gao, Catherine L Smith, Jillian F Ikin, David Brown, Natasha Kinsman, Yuming Guo, Michael J Abramson, Karen Walker-Bone, Malcolm R Sim, Tyler J Lane

## Abstract

**Background:** The link between chronic exposure to ambient PM_2.5_ and lung cancer is well established. However, there is limited evidence on the effects of more acute, high-level exposure such as that resulting from the 2014 Hazelwood coal mine fire in regional Australia. We investigated the effects of PM_2.5_ from the mine fire on cancer incidence 8.5 years later.

**Methodology:** We obtained Victorian Cancer Registry data linked to 2872 Hazelwood Health Study Adult Cohort members, for the period August 2014 to December 2022. Individual fire-related PM_2.5_ exposure was estimated by blending time-location diaries with spatial and temporal air pollution modelling data. To assess the cancer risk associated with fire related PM_2.5_ exposure, we employed competing risk regression models, accounting for people who died from causes other than cancer during follow-up and adjusting for confounders, including cigarette smoking.

**Results:** In the post-mine fire period, 295 people (14.3/1000 person-years) were diagnosed with 332 new cancers (12.7/1000 person-years). No significant association was found between fire-related PM_2.5_ exposure and the overall incidence of cancer (HR = 1.00, 95%CI: 0.90-1.11). Additionally, no associations were identified with any specific cancer subtypes, including lung cancer. However, a higher risk of overall cancer incidence was observed in Morwell compared with Sale, (HR = 1.27, 95% CI: 0.93–1.73).

**Conclusions:** While we found no evidence that this coal mine fire increased cancer incidence, it would be premature to rule out potential carcinogenic effects. Cancer has a long latency period, which means it will be necessary to analyse new data as they become available to more conclusively determine the effects of medium-duration, high-level smoke exposure.

## 1 Introduction

In February 2014, the Hazelwood open-cut coal mine in the Latrobe Valley in regional Victoria, Australia, caught fire and burned for approximately 45 days, shrouding the nearby town of Morwell in smoke and ash. In response to community concerns about potential long-term health effects, the Victorian Department of Health commissioned the Hazelwood Health Study (1–3).

One concerning health consequence was the potential to develop lung cancer. However, at the time there was limited research on the effects of mine fire smoke or wildfire smoke on cancer incidence, and while indoor coal combustion for cooking and heating provided a useful proxy, it may not have may not have fully represented the specific risks associated with coal-mine fire exposure (4). Smoke from the fire contained high concentrations of fine particulate air pollution <2.5 μm in diameter, commonly referred to as PM_2.5_ (5). Though characterised by its small size, PM_2.5_ is a complex mix of different particles and sizes that can have various sources of origin (such as agriculture, industrial combustion, surface transportation, residential and commercial sources, waste disposal and large-scale fires) (6,7). Despite their small size, PM_2.5_ particles can have relatively large surface areas, toxin absorbing capabilities, and the ability to penetrate deep into airways and become absorbed in the bloodstream, causing harm throughout the body (8,9).

Ambient PM_2.5_ is associated with increased risk of lung and urological cancers, with mixed evidence for other cancers like breast cancer (10,11). There are many proposed causal mechanisms, including: activation of oncogenes through alteration of microRNA expression (8,12); gene mutations (8,13); deactivation of tumour suppression genes like p53 through methylation (8,14,15); promotion of angiogenesis, which contributes to metastasis and tumour growth (8); and disruption of autophagy and apoptosis (8,16). There is some evidence that landscape fires may produce more harmful mixtures than ambient sources such as traffic and industrial activities, particularly to people living with cancer (17).

The Hazelwood Health Study has conducted two investigations of the potential effects of mine fire-related PM_2.5_ on cancer. The first was based on Adult Cohort members linked to cancer registry data with five years of post-fire follow-up (18). The second used population-wide anonymous cancer registry data and 8.5 years of follow-up data (19). Neither study found evidence for increased incidence of cancer. However, each had its limitations; the linked analysis had a relatively short follow-up period, while the anonymous extract lacked individual-level information on exposure and potential confounders.

In this analysis, we use 8.5 years of linked cancer data to investigate whether medium-duration exposure to extreme amounts of coal mine fire-related PM_2.5_ was associated with cancer incidence.

## 2 Methods

This study replicates the methods of the Hazelwood Health Study’s first cancer incidence linkage analysis (18). We have applied similar methods in this study, whilst noting any departures in approach.

### 2.1 Participants

Study participants were derived from the Hazelwood Health Study’s Adult Cohort, which was established in 2016/2017. The cohort comprised persons who were aged 18 or older and living in Morwell (exposed) or selected areas in nearby Sale (unexposed comparison) at the time of the mine fire, and who completed the Hazelwood Health Study Adult Survey (2). The analysis examines how 8.5 years of cancer data determines the medium-term exposure to high levels of coal mine fire-related PM_2.5_ contributed to cancer incidence. Out of 4,056 cohort members, n=2,872 (2,223 from Morwell and 649 from Sale) consented to have their responses linked to administrative health datasets including the Victorian Cancer Registry (VCR) (19) and National Death Index (NDI)(2).

### 2.2 Outcomes

We obtained VCR data on new cancer diagnoses from six months post-fire to the most recent available (9 August 2014 to 31 December 2022). Cancers were limited to malignant tumours based on ICD-10 coding (C00-97, D45-D46, D47.1-D47.5). Subtypes based on location included digestive (ICD-10 codes: C15-C25), colorectal (C18-C21), respiratory and intrathoracic (C30-C38), breast (C50), male reproductive organs (C60-C63), urinary tract (C64-C68), and lymphoid, haematopoietic and related tissues (C18-96, D45-46, D47.1, D47.3-D47.5). NDI data to June 2023 was obtained to identify deaths during the follow-up period.

### 2.3 Exposures

Due to limited air quality monitoring throughout the Latrobe Valley, especially in the early, most intense periods of the coal mine fire, the Commonwealth Scientific and Industrial Research Organisation (CSIRO) modelled fire emissions using chemical transport and dispersion models and conducted larger-scale modelling for background concentrations (5). Time-location diaries included in the 2016/17 Adult Survey captured participants’ self-reported whereabouts during the mine fire in 12-hour intervals (2). When combined with modelled PM_2.5_ data, these were converted to daily means throughout the fire period to provide estimates of individual exposure levels (2).

### 2.4 Confounders

Analyses were adjusted for the same potentially confounding factors as in the previous linkage study, which had been collected as part of the Adult Survey (18). These included : age (</≥60 years of age) at the time of the fire, sex, marital status, educational attainment (secondary education to year 10, education to years 11-12, certificate/diploma/tertiary), employment status at the time of the survey (unemployed, unable to work, unpaid occupation and employed), occupational exposures (not exposed, coal mine or power station exposure, other occupational exposures including construction, driving, farming, asbestos, etc.), smoking status (non-smoker, former, current smoker), cigarette pack-years (not used in previous analysis), alcohol consumption (non-drinker, low, high risk), and Index of Relative Socioeconomic Advantage and Disadvantage (IRSAD; not used in the previous analysis) providing a continuous indicator of a residential area socioeconomic status.

### 2.5 Analysis

Descriptive statistics incorporated participant demographics, confounders, and cancer outcomes, to enable comparison between the exposed (Morwell) and unexposed participants (Sale). Comparisons were performed using Kruskal-Wallis tests for continuous variables, presented as Mean (SD), and Fisher’s exact test for dichotomous and categorical variables. Changes in post-fire cancer incidence were evaluated using the competing risk regression models *tidycmprsk* package (20) in R using RStudio (21,22). These models estimate hazard ratios based on the subdistribution hazard function which represents the immediate risk for the development of cancer for each subtypes, while accounting for the competing risk of death (23,24). This approach ensures that the analysis appropriately adjusts for individuals who may have died before a cancer diagnosis, providing a more accurate assessment of cancer risk. The primary outcome was cancer diagnosis, with the competing event defined as death from any cause other than cancer (25). Results are reported as Hazard Ratios (HRs) and 95% confidence intervals (CIs). To account for missing data, we applied multiple imputation with random forests (26), with the number of imputations equivalent to the proportion of cases with missing data, and pooled results according to Rubin’s rules (27).

### 2.6 Ethics

This study is a part of the Hazelwood Adult Survey & Health Record Linkage Study (Project ID: 25680; previously CF15/872-2015000389).

## 3 Results

### 3.1 Description of cohort

The post-fire period (9 August 2014 to 31 December 2022) consisted of 23,273 persons years (Morwell: 18,014; Sale: 5,259) when accounting for people who died during follow-up. There were 332 newly diagnosed cancers (Morwell: 269; Sale: 63) among 295 individuals (Morwell: 241; Sale: 54). The incidence was 14.3 per 1,000 person years (Morwell: 14.9; Sale: 12.0) and the prevalence of individuals diagnosed with cancer during the follow-up period was 12.7 cancers per 1,000 person years (Morwell: 13.4; Sale: 12.0), both of which were slightly higher in Morwell. Comparing study sites in the post-fire period indicated Morwell had an elevated but non-significantly different incidence of cancer, whether in crude (HR: 1.30, 95%CI: 0.97-1.75) or adjusted models (HR: 1.27, 95%CI: 0.93-1.73); there were no detectable differences by cancer type, though male reproductive organ cancers were elevated in both the crude (HR: 2.41, 95%CI: 0.95-6.09) and adjusted models (HR: 2.32, 95%CI: 2.32, 95%CI: 0.88-6.16). Comparisons between Morwell and Sale can be found in Table S1.

There were some demographic differences between exposed and unexposed participants, which are presented in Table 1. Morwell participants had less educational attainment, fewer people in paid employment and lower residential socioeconomic status. Occupational exposures related to coal mines and power stations were higher in Morwell, which was unsurprising as it was the site of a mine and power station (both closed in 2017), whereas other occupational exposures (e.g. construction, driving, farming, asbestos removal, etc.) were higher in Sale. Sale participants also had a higher proportion of high-risk drinking behaviour.

**Table 1:** Description of cohort.

|  | Morwell, N = 2,223 | Sale, N = 649 | p-value |
| --- | --- | --- | --- |
| **PM<sub>2.5</sub> exposure, Mean (SD)** | **15.3 (11.7)** | **0.0 (0.3)** | **< 0.001** |
| **Deceased, n (%)** | 224 (10%) | 69 (11%) | 0.712 |
| **Female, n (%)** | 1,196 (54%) | 372 (57%) | 0.117 |
| **Age 60+ years, n (%)** | 975 (44%) | 287 (44%) | 0.893 |
| **Married/de facto, n (%)** | 1,353 (61%) | 422 (65%) | 0.060 |
| **Educational attainment, n (%)** |  |  | **< 0.001** |
| Secondary up to year 10 | **708 (32%)** | **148 (23%)** |  |
| Secondary year 11-12 | **439 (20%)** | **107 (17%)** |  |
| Certificate/Diploma/Tertiary | **1,062 (48%)** | **391 (61%)** |  |
| Missing | **14** | **3** |  |
| **Employment, n (%)** |  |  | **< 0.001** |
| Employed | **916 (41%)** | **293 (45%)** |  |
| Unpaid Occupation | **1,010 (46%)** | **312 (48%)** |  |
| Unemployed | **105 (4.8%)** | **12 (1.9%)** |  |
| Unable to Work | **177 (8.0%)** | **30 (4.6%)** |  |
| Missing | **15** | **2** |  |
| **Occupational exposure, n (%)** |  |  | **< 0.001** |
| Not exposed | **1,251 (56%)** | **409 (63%)** |  |
| Coal mine/power station exposed | **379 (17%)** | **21 (3.2%)** |  |
| Other occupational exposures | **593 (27%)** | **219 (34%)** |  |
| **Pack years (current and former smokers only), Mean (SD)** | 20.1 (20.2) | 20.8 (22.6) | 0.711 |
| Missing | 25 | 25 |  |
| **Smoking status, n (%)** |  |  | **0.009** |
| Non-smoker | **1,028 (47%)** | **341 (53%)** |  |
| Former smoker | **802 (36%)** | **215 (33%)** |  |
| Current smoker | **379 (17%)** | **87 (14%)** |  |
| Missing | **14** | **6** |  |
| **Drinking risk (AUDIT-C score), n (%)** |  |  | **0.010** |
| Non-drinker | **548 (25%)** | **150 (23%)** |  |
| Low risk | **798 (37%)** | **206 (32%)** |  |
| High risk | **815 (38%)** | **284 (44%)** |  |
| Missing | **62** | **9** |  |
| **IRSAD (Socioeconomic area) score, Mean (SD)** | **859.2 (94.0)** | **908.0 (51.2)** | **<0.001** |

### 3.2 Fire-related PM_2.5_ and cancer incidence

Crude and adjusted hazard ratios for the associations between cancer incidence and 10µg/m^3^ increments in fire-related PM_2.5_ are presented in Figure 1 and Table S2. There were no detectable associations between fire-related PM_2.5_ exposure and overall cancer incidence (adjusted HR: 1.00, 95%CI: 0.90-1.11), nor were there any detectable associations within cancer subtypes. The cancer subtypes with the highest point estimates were urinary tract, respiratory and intrathoracic, but due to small numbers, these risk estimates were imprecise as indicated by the wide confidence intervals (see Figure 1 and Table S1).

**Figure 1.**
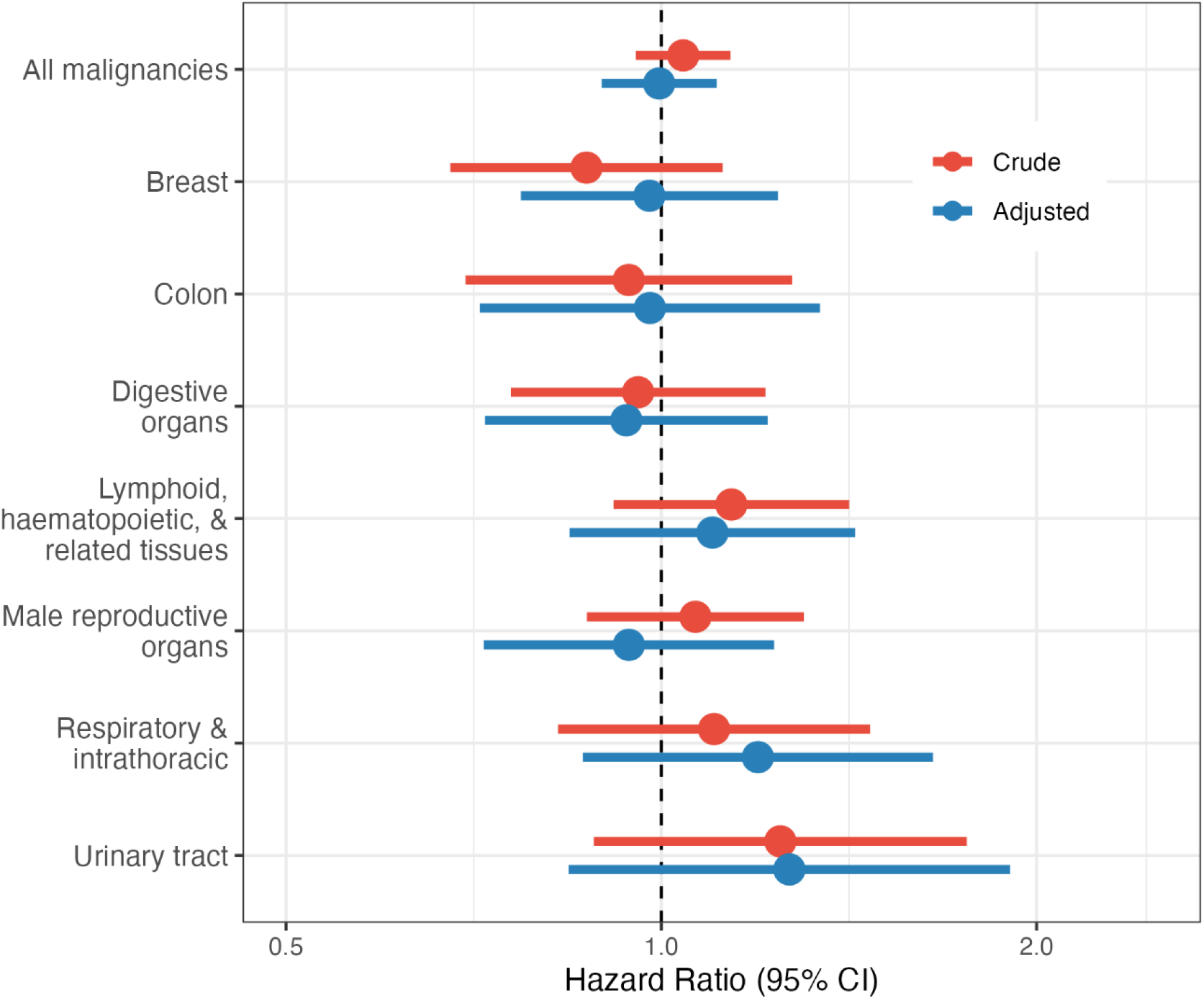
Forest plot showing crude and adjusted Hazard Ratios for incidence of all cancers, and cancer subtypes, per 10-μg/m^3^ increase in mine-fire related PM_2.5_ exposure, using a competing risks model adjusting for sex, age, marital status, education, employment status, smoking and drinking behaviour, occupational exposures and town.

## 4 Discussion

We investigated the effects of coal mine fire-related PM_2.5_ exposure on cancer incidence over 8.5 years following the Hazelwood coal mine fire, among 2872 Hazelwood Health Study Adult Cohort members. While post-fire cancer incidence and prevalence were marginally higher among Morwell compared with Sale participants, these may reflect previously elevated rates of some cancer subtypes in the local area and possibly be attributed to risk factors including occupational exposures and socioeconomic and lifestyle factors (28,29). However, we found no evidence that cancer incidence was associated with fire-related PM_2.5_ exposure. These results align with those of our previous 5-year linkage study (16) but, importantly, adding several additional years of follow up data.

It is worth examining why our findings to date might differ from previous evidence about the carcinogenic effects of ambient PM_2.5_, which has been consistently linked to increased cancer incidence (10,11). One possible explanation is differences in duration of exposure. The effect of ambient air pollution on cancer incidence can take at least a decade to manifest (30). Acute exposures may be similar. Cancer latency is another key factor. The lag between chronic air pollution exposure and cancer onset can take decades (30). For instance, it was more than a decade before there was evidence that first responders to the 9/11 World Trade Center terrorist attacks had elevated risk of certain cancers (31,32). As a result, studies with shorter follow-up such ours may not capture the periods when new exposure-related cancers emerge in detectable numbers. Moreover, the smaller population and sample size in Morwell, compared to studies in densely populated areas such as New York City, constrains the statistical power needed to detect an effect. However, despite these limitations, smaller studies remain important, particularly when aggregated with similar research across multiple disaster events.

It is also possible that smoke from the Hazelwood mine fire, while shown to harm health in other ways, may not have been sufficient to affect cancer incidence rates, at least in ways that can be detectable with our sample size or within this time frame (33–36). In other analyses with similar follow-up but a larger sample size using anonymous cancer registry data, we also found no evidence of an increase in cancer associated with PM_2.5_ exposure (37). However, this larger analysis relied on population-wide anonymous extracts, where exposure to fire-related PM_2.5_ was determined based on area of residence at time of cancer diagnosis, not during the mine fire, meaning there was an unknown degree of misclassification.

Our analyses were also impeded by external events. During the COVID-19 pandemic-related lockdowns, new cancer diagnoses fell by about 10% across Victoria (38). While the effect was smaller in regional and socioeconomically deprived areas, it was still detectable in our previous analysis of anonymous cancer registry data (37). The effect would have been to reduce statistical power at the latter end of our study period when, due to its long latency, we would expect to see the most difference in cancer diagnoses if the mine fire had an effect. This meant that, in addition to the lag between carcinogen exposure and cancer onset, there may have been a lag between cancer onset and detection/diagnosis in regional and socioeconomically deprived areas as a result of the pandemic.

In short, while we have yet to find evidence that the Hazelwood coal mine fire increased cancer incidence, it would be premature, given the relatively short follow-up time, to conclude that it has not had an effect. Longer follow-up times are required to arrive at firmer conclusions.

### 4.1 Strengths and Limitations

This study has several strengths. Our Adult Survey data captured important information to construct a measure of individual-level exposures as well as important confounders such as socioeconomic status and tobacco and alcohol use. The use of competing risk regressions accounted for non-cancer deaths, and multiple imputation accounted for missing data, both increasing statistical power and adjusting for any bias due to any missingness at random.

Nevertheless, there were several limitations. The relatively short follow-up period and the small sample size constrained our ability to detect significant changes, particularly in rarer cancers. These limitations should be considered when interpreting the findings and their implications for cancer incidence. The effects of the COVID-19 pandemic and associated restrictions have potentially made it more difficult at present to detect changes in cancer incidence caused by the mine fire.

## 5 Conclusions

We did not find evidence that smoke-related PM_2.5_ from the Hazelwood coal mine fire increased incidence of cancer. However, considering the long latency period of cancer and delays in diagnoses, particularly in regional areas with limited access to health services, it would be premature to rule out potential carcinogenic effects. It would be wise to continue monitoring cancer data in this cohort over a longer follow-up period in order to arrive at firmer conclusions.

## Data Availability

All data produced in the present study are available upon reasonable request to the authors.

## 6 Supplementary Materials

**Table S1:**
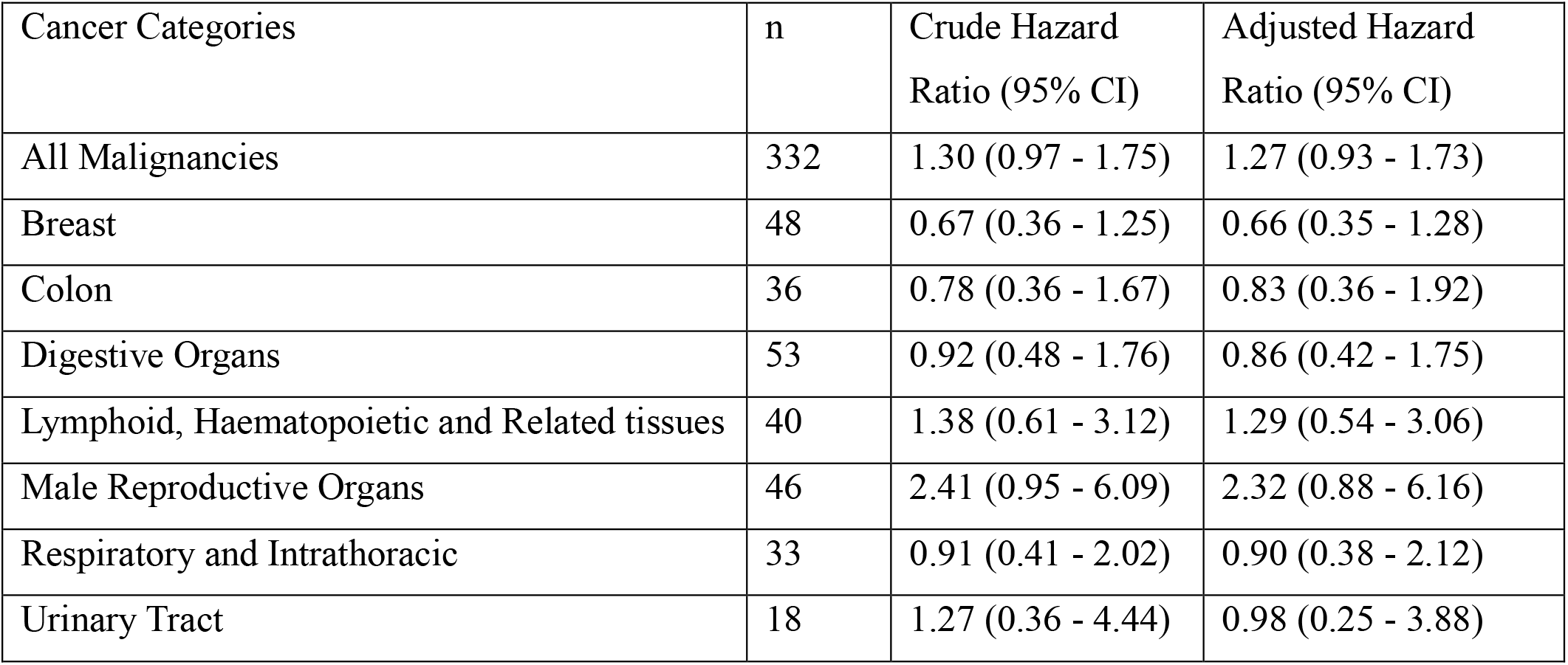
Crude and adjusted Hazard Ratios for incidence of all cancers, and cancer subtypes, comparing Morwell to Sale.

| Cancer Categories | n | Crude Hazard Ratio (95% CI) | Adjusted Hazard Ratio (95% CI) |
| --- | --- | --- | --- |
| All Malignancies | 332 | 1.30 (0.97 - 1.75) | 1.27 (0.93 - 1.73) |
| Breast | 48 | 0.67 (0.36 - 1.25) | 0.66 (0.35 - 1.28) |
| Colon | 36 | 0.78 (0.36 - 1.67) | 0.83 (0.36 - 1.92) |
| Digestive Organs | 53 | 0.92 (0.48 - 1.76) | 0.86 (0.42 - 1.75) |
| Lymphoid, Haematopoietic and Related tissues | 40 | 1.38 (0.61 - 3.12) | 1.29 (0.54 - 3.06) |
| Male Reproductive Organs | 46 | 2.41 (0.95 - 6.09) | 2.32 (0.88 - 6.16) |
| Respiratory and Intrathoracic | 33 | 0.91 (0.41 - 2.02) | 0.90 (0.38 - 2.12) |
| Urinary Tract | 18 | 1.27 (0.36 - 4.44) | 0.98 (0.25 - 3.88) |

**Table S2:** Crude and adjusted Hazard Ratios for incidence of all cancers, and cancer subtypes, per 10 μg/m^3^ increase in mine-fire related PM_2.5_ exposure.

| Cancer Categories | n | Crude Hazard Ratio (95% CI) | Adjusted Hazard Ratio (95% CI) |
| --- | --- | --- | --- |
| All Malignancies | 332 | 1.04 (0.95 - 1.14) | 1.00 (0.90 - 1.11) |
| Breast | 48 | 0.87 (0.68 - 1.12) | 0.98 (0.77 - 1.24) |
| Colon | 36 | 0.94 (0.70 - 1.27) | 0.98 (0.72 - 1.34) |
| Digestive Organs | 53 | 0.96 (0.76 - 1.21) | 0.94 (0.72 - 1.22) |
| Lymphoid, Haematopoietic and Related tissues | 40 | 1.14 (0.92 - 1.41) | 1.10 (0.84 - 1.43) |
| Male Reproductive Organs | 46 | 1.07 (0.87 - 1.30) | 0.94 (0.72 - 1.23) |
| Respiratory and Intrathoracic | 33 | 1.10 (0.83 - 1.47) | 1.20 (0.87 - 1.65) |
| Urinary Tract | 18 | 1.25 (0.88 - 1.76) | 1.27 (0.84 - 1.90) |

